# Screening for high amounts of SARS-CoV-2 identifies pre-symptomatic subjects among healthy healthcare workers

**DOI:** 10.1101/2020.12.13.20248122

**Authors:** Joakim Dillner, K. Miriam Elfström, Jonas Blomqvist, Lars Engstrand, Mathias Uhlén, Carina Eklund, Fredrik Boulund, Camilla Lagheden, Marica Hamsten, Sara Nordqvist-Kleppe, Maike Seifert, Cecilia Hellström, Jennie Olofsson, Eni Andersson, August Jernbom Falk, Sofia Bergström, Emilie Hultin, Elisa Pin, Ville N. Pimenoff, Sadaf Hassan, Anna Månberg, Peter Nilsson, My Hedhammar, Sophia Hober, Johan Mattsson, Laila Sara Arroyo Mühr, Kalle Conneryd Lundgren

## Abstract

**Background:** Pre-symptomatic subjects are spreaders of SARS-CoV-2 infection, and strategies that could identify these subjects, particularly in hospital settings, are needed.

**Methods:** We tested a cohort of 9449 employees at work at the Karolinska University Hospital, Stockholm, Sweden for SARS-CoV-2 RNA and antibodies, linked the screening results to sick leave records and examined the association between screening results and past or future sick leave using multinomial logistic regression.

**Results:** We found that healthcare workers with high amounts of SARS-CoV-2 virus, as indicated by the Cycle threshold (Ct) value in the PCR, had the highest risk for sick leave in the two weeks <u>after</u> testing (OR 11·97 (CI 95% 6·29-22·80)) whereas subjects with low amounts of virus had the highest risk for sick leave in the past three weeks <u>before</u> testing (OR 6·31 (4·38-9·08)). Only 2·5% of employees were SARS-CoV-2 positive while 10·5% were positive by serology and 1·2% were positive in both tests. Serology-positive subjects were not at excess risk for future sick leave (OR 1·06 (95% CI, 0·71-1·57)), but virus-positive subjects had a 7·23 fold (95% CI, 4·52-11·57)) increased risk for sick leave within two weeks post testing.

**Conclusions:** Screening of asymptomatic healthcare workers for high amounts of SARS-CoV-2 virus using Ct values will identify pre-symptomatic subjects who will develop disease in the next few weeks. Identification of potentially contagious, pre-symptomatic subjects is likely critical for protecting patients and healthcare workers.

**Main point:** Healthy healthcare workers with low amounts of SARS-CoV-2 nucleic acids will previously have had the disease. Presence of a high amount of SARS-CoV-2 nucleic acids predicts future symptomatic disease.

## Introduction

The current epidemic of SARS-CoV-2 is largely driven by asymptomatic individuals [1, 2]. To design strategies for SARS-CoV-2 control, effective identification of infectious subjects in defined communities is critical [3]. The incubation time from exposure to onset of symptoms has been estimated to last a median of six days [1], with peak infectiousness occurring zero to two days before onset of symptoms and pre-symptomatic spread estimated to account for a substantial proportion of disease transmission [1, 2]. While infectiousness decreases with increasing time after onset of symptoms, viral nucleic acids can still be detected after resolution of symptoms, in one study even six weeks after symptom resolution [4, 5]. If the virus is still present after symptom resolution it is usually only in low amounts and appears to not be viable [4, 5]. Screen-detected positivity may mark subjects who are symptomatic, pre-symptomatic (will develop symptoms later), post-symptomatic (symptoms have resolved), or asymptomatic (will never develop symptomatic disease) [6].

To identify potentially contagious subjects among asymptomatic healthcare workers (HCWs) is particularly important for SARS-CoV-2 control as various healthcare-related outbreaks have been observed) [3, 7, 8]. Knowledge of the extent of spread enables assessment of the infectious disease control in the healthcare setting, which is important both for adequate staffing in a critically important sector of society and for continued public trust and adequate healthcare-seeking behavior [9].

Antibodies to SARS-CoV-2 develop rather slowly, commonly concomitantly with symptom resolution and increases in subsequent weeks [10]. The PCR test, in addition to providing a dichotomous positive or negative result, will also provide a semiquantitative measure of the amount of virus present, the Cycle Threshold value (Ct) [5]. The Ct value is the number of sample amplification cycles needed before the virus was detectable. For example, a sample with a Ct value of 3 contains >30 billion times more virus than a sample that is positive with a Ct value of 38 and the case has been made that contact tracing should focus on the subjects with a large amount of virus [5].

It is particularly important to obtain data on how the SARS CoV-2 testing results relate to pre-symptomatic disease or post-symptomatic disease in a manner that is free from recall bias. To address this, we invited all employees currently on duty at the Karolinska University Hospital, Stockholm, Sweden to participate in a study that concomitantly measured presence of SARS-CoV-2 viral nucleic acid in throat samples and presence of antibodies to the virus in serum samples, in relation to the sick leave records of the participants.

## Materials and Method

The Karolinska University Hospital has about 15,300 employees. The hospital announced that all HCWs on duty were welcome to participate in a study that evaluated the concomitant presence of viral nucleic acids in throat swabs and presence of antibodies to the virus in serum. Participants were recruited between April 23^rd^, 2020 and June 24^th^, 2020. All enrolled participants signed a written informed consent that also included permission to extract data from the employer’s administrative databases that included data on sick leave. The study was approved by the National Ethical Review Agency of Sweden (Decision number 2020-01620). Trial registration number: ClinicalTrials.gov NCT04411576.

### Viral Nucleic Acid Detection

Throat swab samples were obtained using the Beaver Specimen Collection kit (<u>stratech.co.uk/wp-content/uploads/2020/04/BEAVER-IFU-43903-Sample-Collection-Kit19324.pdf</u>) as described in the users’ manual. Sample preparation followed safety routines according to BSL2 requirements including negative pressure in the room, biosafety cabinets, and installed HEPA filters. Samples were heat inactivated for 50 minutes at 75 degrees C. Extraction of viral RNA was performed using the MGISP-960 automated extraction standard workflow, according to the manufacturer’s protocol (Wuhan MGI Tech Co, Ltd) using the MGIEasy Magnetic Beads Virus DNA/RNA extraction kit. The BGI 2019-nCov Detection kit (BGI Real-Time RT-PCR for detecting 2019 nCoV) was used according to the manufacturer’s instructions, including internal parameters to monitor sampling quality and testing process. RT-PCR was performed on QuantStudio5 instruments and software (Design and Analysis Software v1.5.1, Thermo Scientific). All steps in the diagnostic pipeline followed standard operating protocol validated for reproducibility, sensitivity, and specificity, including lack of cross-reactivity with other Coronavirus strains.

### Serological analyses of antibodies

Whole blood was collected in serum-separating tubes and centrifuged at 2000 x g for 10 minutes. Serum samples were inactivated by heat-treatment at 56 degrees C for 30 minutes and then stored at −20 degrees C until further analysis.

Serological reactivity was measured towards three different virus protein variants, (i) Spike trimers comprising the prefusion-stabilized spike glycoprotein ectodomain [11] expressed in HEK-cells and purified using a C-terminal Strep II tag), (ii) Spike S1 domain, expressed in CHO-cells and purified using C-terminal HPC4-tag, and (iii) Nucleocapsid protein, expressed in *E*.*coli* and purified using a C-terminal His-tag. The sera were analyzed using a multiplex antigen bead array in a 384-plate format using a FlexMap3D instrument (Luminex Corp) with IgG detection [12]. The serology assay was then evaluated based on the analyses of 154 samples from Covid-19 subjects (defined as PCR-positive individuals sampled more than 16 days after disease onset) and 321 negative samples (defined as samples collected 2019 or earlier in the same region, including 26 individuals with confirmed infections of other Coronaviruses than SARS-CoV-2). The assay had a 99.4% sensitivity and 99.1% specificity. The cut-off for seropositivity was defined for each antigen as mean +6SD of 12 negative control samples included in each analysis batch. To be assigned as IgG positive, a sample was required to show reactivity against at least two of the three included viral antigens. Serum IgG bound to antigen coated beads was detected by fluorescent anti-hIgG (Invitrogen, H10104) and recorded as relative fluorescence intensity (AU).

### Data analyses

Screening test results were examined separately and as a combined categorical variable. PCR positivity was dichotomized into strongly (<27·0) and weakly positive (greater than or equal to 27·0) based on the median Ct value among PCR positive/serology negative participants, rounded down to the nearest whole integer. Antibody positivity modified the risk for sick leave associated with PCR positivity (p= 0·0008). Therefore, a combined variable of PCR and serology results was used to examine the association between SARS-CoV-2 status and sick leave. A combined variable with four categories was used to simultaneously examine serology and PCR test results. Descriptive statistics were used to examine test results by age and sick leave. A multinomial logistic regression examined the association between test results and sick leave measured as a categorical variable, adjusted for age in 10-year categories and sex. Sick leave in the six weeks prior to testing and two weeks after testing was categorized as either no sick leave during the period of interest (reference category), sick leave in the 4-6 weeks before testing, sick leave in the 1-3 weeks before testing and sick leave in the two weeks after testing. For subjects with sick leave in more than one category, the period with the highest number of sick leave days was chosen. If two periods had an equal number of sick leave days, the period further back in time was chosen. With conventional statistical power and confidence while assuming a cumulative proportion of sick leave among non-exposed persons of 30% and that 10% of the cohort might be exposed, about 3,800 subjects would need to be enrolled to be able to detect associations of 1.4 or greater. Analyses used SAS 9.4, Cary, NC.

## Results

The Karolinska University Hospital had approximately 15,300 employees in the spring of 2020. Of these, 14,201 were enrolled in this study. After exclusion of HCWs not formally employed (e.g. medical students) and those without valid results on both the PCR and serology tests, the final cohort consisted of 9,449 subjects with complete data on sick leave and valid results on both tests (Figure 1), well over the estimated number needed for sufficient statistical power.

**Figure 1.**
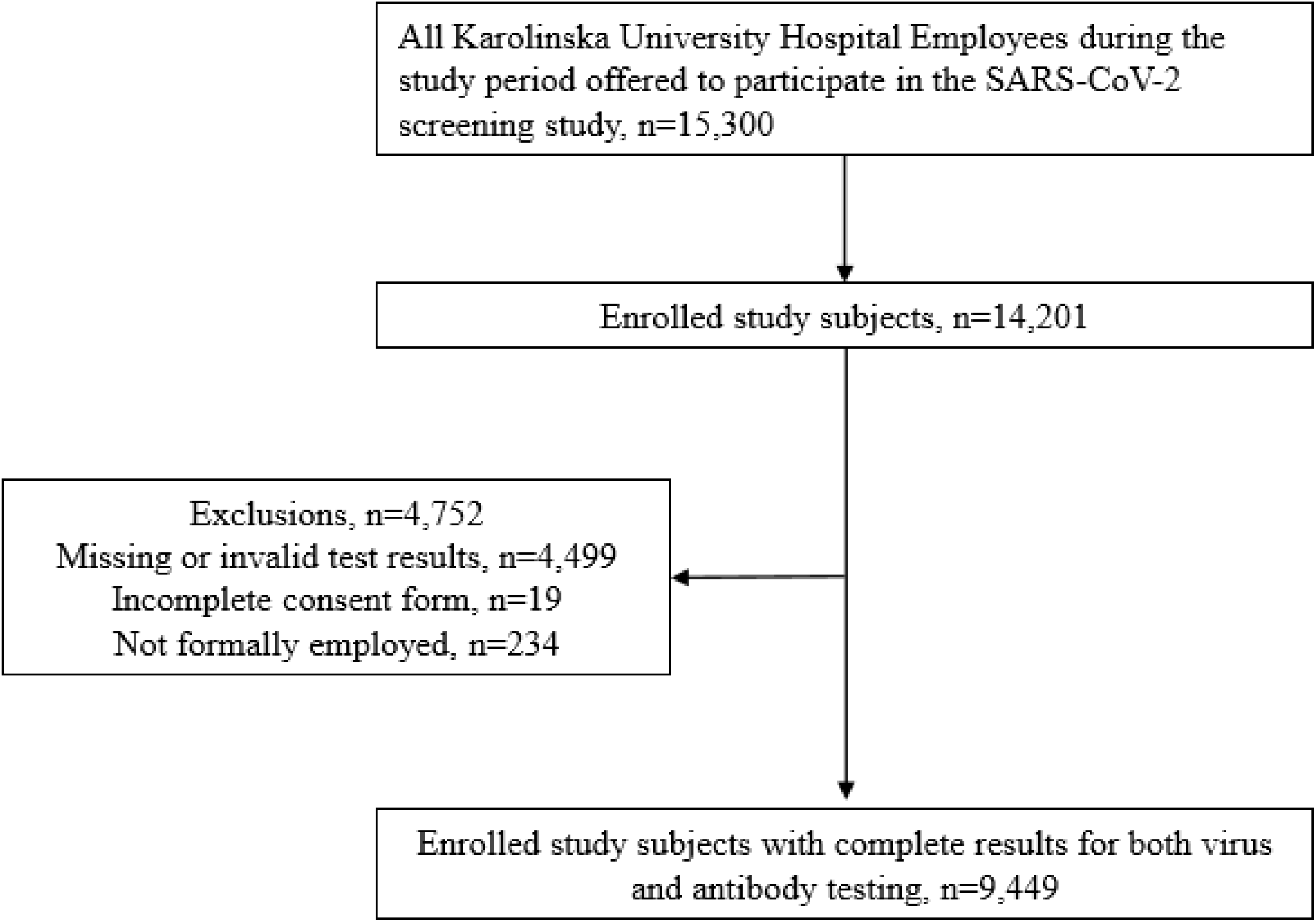
STROBE flowchart of study subjects.

The overall number and proportion of employees that tested positive or negative in the two tests are shown by age in 10-year spans in Table 1. Seropositivity was most common in the youngest age group (14·9% were positive among subjects under the age of 29) and decreased with age (p-value for trend <0·0001). In total, 88·2% (87·5-88·9) of subjects were negative on both tests, 9·3% (8·7-9·9) were serology positive only, 1·3% (1·1-1·5) were positive for the virus, and 1·2% (1·0-1·5) were positive for both antibodies and the SARS-CoV-2 virus.

**Table 1.** Detection of SARS CoV-2 virus and antibodies to the virus among 9,449 employees of the Karolinska University Hospital, by age.

|  |  |  |  |  |  |  |
| --- | --- | --- | --- | --- | --- | --- |
| Age | Serology positive<br>n (%)* | PCR negative &<br>serology negative<br>n (%) | PCR negative &<br>serology positive<br>n (%) | PCR positive &<br>serology negative<br>n (%) | PCR positive &<br>serology positive<br>n (%) | Total |
| <29 | 170 (14.9) | 956 (83.6) | 157 (13.7) | 17 (1.5) | 13 (1.1) | 1,143 |
| 30-39 | 259 (11.0) | 2,055 (87.6) | 217 (9.3) | 33 (1.4) | 42 (1.8) | 2,347 |
| 40-49 | 251 (10.8) | 2050 (87.8) | 219 (9.4) | 33 (1.4) | 32 (1.4) | 2,334 |
| 50-59 | 209 (9.4) | 1,985 (89.6) | 188 (8.5) | 21 (1.0) | 21 (1.0) | 2,215 |
| 60+ | 106 (1.1) | 1,289 (91.4) | 98 (7.0) | 15 (1.1) | 8 (0.6) | 1,410 |
| Total (n, % (95%<br>CI) | 995, 10.5 (9.9-11.2) | 8,335, 88.2 (87.5-88.9) | 879, 9.3 (8.7-9.9) | 119, 1.3 (1.1-1.5) | 116, 1.2 (1.0-1.5) | 9,449 |

\*Serology positive, regardless of PCR result, Cochran-Armitage Trend Test p-value <0.0001

Overall, 54·5% (38·3% in weeks 1-3 before testing and 16·2% in weeks 4-6) of PCR-positive subjects had a history of sick leave during the past six weeks (post-symptomatic), whereas 63% of seropositive subjects had such history (29·3% in weeks 1-3 before testing and 33·7% in weeks 4-6) (Table 2). In the two weeks after testing, 15·3% of PCR positive subjects had sick leave reported, compared to only 3·4% of the seropositives. By comparison, 5·2% of the double negative subjects had sick leave after the sampling. Among PCR positive subjects, 30·2% did not have any sick leave recorded, neither before nor after testing (asymptomatic).

**Table 2.** Distribution of background characteristics and screening results, by sick leave.

|  |  |  |  |  |  |
| --- | --- | --- | --- | --- | --- |
|  | n, % (95% CI) | Sick leave |  |  |  |
|  |  | No sick leave<br>n (%) | 1-2 weeks<br>after testing<br>n (%) | 1-3 weeks<br>before testing<br>n (%) | 4-6 weeks<br>before testing<br>n (%) |
| Age |  |  |  |  |  |
| 20-29 | 1,143 (12.1) | 648 (56.7) | 77 (6.7) | 180 (15.8) | 238 (20.8) |
| 30-39 | 2,347 (24.8) | 1,270 (54.1) | 162 (6.9) | 376 (16.0) | 539 (23.0) |
| 40-49 | 2,334 (24.7) | 1,403 (60.1) | 104 (4.5) | 344 (14.7) | 483 (20.7) |
| 50-59 | 2,215 (23.4) | 1,359 (61.4) | 105 (4.7) | 296 (13.4) | 455 (20.5) |
| 60+ | 1,410 (14.9) | 934 (66.2) | 52 (3.7) | 160 (11.4) | 264 (18.7) |
| Sex |  |  |  |  |  |
| Female | 7,488 (79.3) | 4,252 (56.8) | 420 (5.6) | 1,156 (15.4) | 1,660 (22.2) |
| Male | 1,961 (20.8) | 1,362 (69.5) | 80 (4.1) | 200 (10.2) | 319 (16.3) |
| SARS-CoV-2 test results |  |  |  |  |  |
| PCR neg/Serology neg | 8,335 (88.2) | 5,230 (62.8) | 436 (5.2) | 1,046 (12.6) | 1,623 (19.5) |
| PCR neg/Serology pos | 879 (9.3) | 313 (35.6) | 28 (3.2) | 220 (25.0) | 318 (36.2) |
| PCR pos/Serology neg | 119 (1.3) | 49 (41.2) | 30 (25.2) | 19 (16.0) | 21 (17.7) |
| PCR pos/Serology pos | 116 (1.2) | 22 (19.0) | 6 (5.2) | 71 (61.2) | 17 (14.7) |
| SARS-CoV-2 PCR test results |  |  |  |  |  |
| PCR negative | 9,214 (97.5) | 5,543 (60.2) | 464 (5.0) | 1,266 (13.7) | 1,941 (21.1) |
| PCR positive | 235 (2.5) | 71 (30.2) | 36 (15.3) | 90 (38.3) | 38 (16.2) |
| PCR weakly positive | 168 (1.8) | 52 (31.0) | 16 (9.5) | 74 (44.1) | 26 (15.5) |
| PCR strongly positive | 67 (0.7) | 19 (28.4) | 20 (29.9) | 16 (23.9) | 12 (17.9) |
| SARS-CoV-2 serology test results |  |  |  |  |  |
| Serology negative | 8,485 (89.5) | 5,279 (62.4) | 466 (5.5) | 1,065 (12.6) | 1,644 (19.5) |
| Serology positive | 995 (10.5) | 335 (33.7) | 34 (3.4) | 291 (29.3) | 335 (33.7) |

Positivity in serology was significantly associated with past history of sick leave (Table 3) but did not confer any risk for future sick leave for the coming two weeks after testing (OR 1·06 (95% CI, 0·71 to 1·57)) (Table 2). By contrast, subjects with viral nucleic acids in the absence of antibodies had very little excess risk for past sick leave (similar to the sick leave history of test-negative subjects), but had a strongly increased risk for imminent sick leave in the two weeks after testing (OR 7·23 (95% CI 4·52-11·57)) (Table 3). Positivity for both virus and for antibodies tended to be most strongly associated with sick leave during the past three weeks (OR 16·51 (95% CI 10·13-26·90)) (Table 3). Compared to PCR negative subjects, strong PCR positivity was in particular associated with future sick leave (OR 11·97 (95% CI 6·29-22·80)) while weak PCR positivity was most strongly associated with sick leave in the 1-3 weeks before testing (OR 6·31 (95% CI 4·38-9·08)). In the multivariate model, male sex and over 50 years of age were associated with lower risk for sick leave (data not shown). The mutual adjustments in the multivariate model had only minor effects on the estimates (Supplementary Table 1). Examining test results and sick leave week by week in the six weeks before test and two weeks after testing highlights these patterns. Sick leave peaked for subjects that tested PCR positive/serology positive two weeks before testing while sick leave increased in the two weeks after testing for PCR positive/serology negative subjects (Figure 2). The amounts of virus as defined by the Ct values revealed a clear pattern of sick leave <u>prior to</u> testing for subjects with low amounts of virus and a sharp increase in sick leave <u>after</u> testing for subjects with a high amount of virus (Figure 3).

**Table 3.** Association between testing results and sick leave*.

| **Table 3. Association between testing results and sick leave\*** |  |  |  |
| --- | --- | --- | --- |
|  | **1-2 weeks after**<br>**testing**<br>**vs No sick leave**<br>**OR (95% CI)** | **1-3 weeks before**<br>**testing**<br>**vs No sick leave**<br>**OR (95% CI)** | **4-6 weeks before**<br>**testing**<br>**vs No sick leave**<br>**OR (95% CI)** |
| **SARS-CoV-2 test results** |  |  |  |
| PCR neg/Serology neg | 1.00 | 1.00 | 1.00 |
| PCR neg/Serology pos | 1.06 (0.71-1.57) | 3.52 (2.92-4.25) | 3.31 (2.80-3.91) |
| PCR pos/Serology neg | 7.23 (4.52-11.57) | 1.92 (1.12-3.28) | 1.38 (0.82-2.31) |
| PCR pos/Serology pos | 3.24 (1.30-8.06) | 16.51 (10.13-26.90) | 2.54 (1.34-4.81) |
| **SARS-CoV-2 PCR test results** |  |  |  |
| PCR negative | 1.00 | 1.00 | 1.00 |
| PCR weakly positive | 3.61 (2.04-6.40) | 6.31 (4.38-9.08) | 1.45 (0.90-2.34) |
| PCR strongly positive | 11.97 (6.29-22.80) | 3.61 (1.84-7.09) | 1.80 (0.87-3.73) |

\*Model 1 with testing results in four categories, adjusted for age and sex. Model 2 with PCR test results in three categories, adjusted for age and sex.

**Figure 2.**
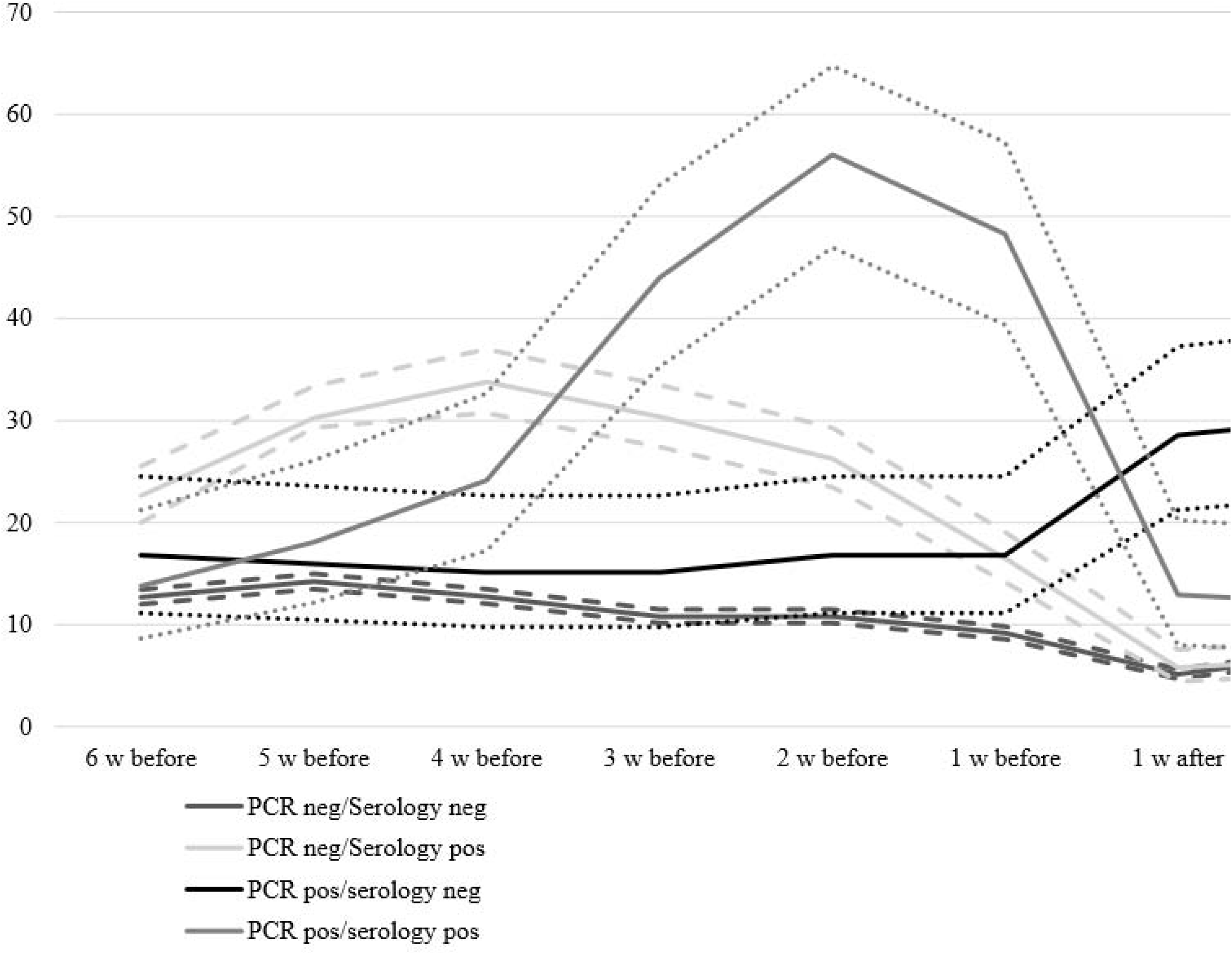
SARS-CoV-2 screening result by sick leave before and after testing.

**Figure 3.**
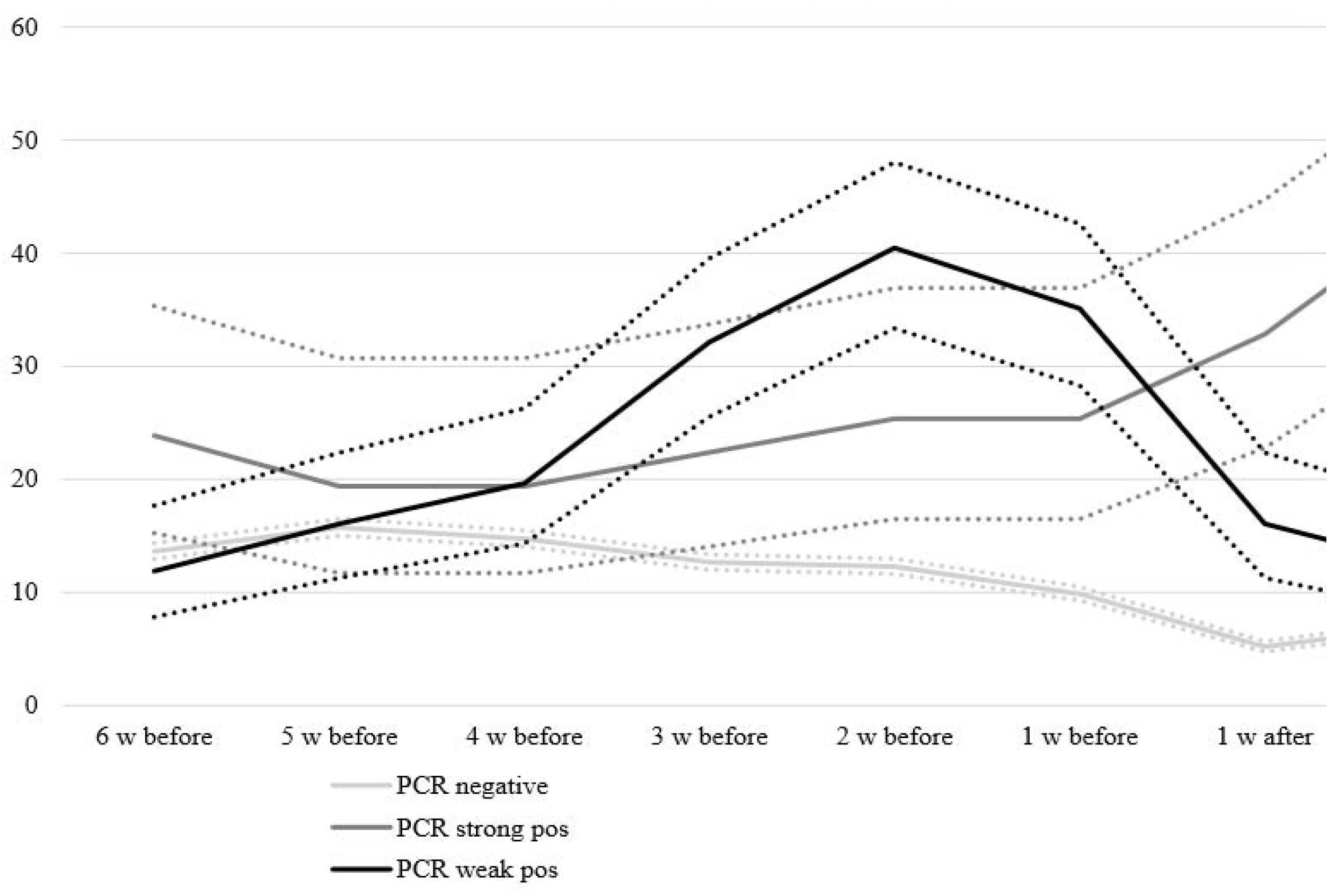
Sick leave among healthcare workers by PCR screening result, stratified b strongly or weakly positive. Dotted lines denote the upper and lower bounds of the 95% confidence limits.

## Discussion

We found that large-scale screening of asymptomatic HCWs identified a limited number of SARS-CoV-2-positive subjects (235/9449 subjects). Among these, more than half were only positive for low amounts of virus (had high Ct values) and had mainly already had disease (post-symptomatic subjects). Systematic reviews have not identified any reports of shedding of live virus for more than 9 days after debut of symptoms, whereas low amounts of virus may be detectable for many weeks after resolution of symptoms [13]. Our large-scale study confirms that the amounts of virus (the Ct value) is useful for distinguishing between post-symptomatic and pre-symptomatic subjects that may be a risk group for transmission and identification of these may be critical for protecting patients and HCWs.

As it is established that subjects who have recovered from symptomatic disease are no longer infectious, it seems appropriate to focus infection control on the subjects with the pattern of pre-symptomatic disease. Positivity in serology is also primarily associated with past sick leave and presence of antibodies may also be useful to identify the subjects with post-symptomatic positivity.

Many studies have reported screening of HCWs as part of an infection control strategy [7, 14]. For example, 3% of HCWs were PCR positive in a major London hospital [15]. Another study reported on an outbreak in a skilled nursing facility, where a large proportion of HCWs tested positive.^7^ Although many studies have screened HCWs with PCR, the amount of virus (the cycle threshold value) has not been taken into account, although this value is obtained when a real-time PCR reaction is performed.

Studies comparing detection of viral nucleic acids and antibodies have mostly been focusing on COVID-19 patients, whereas not on combined PCR/serology screening of healthy workers. This enabled us to provide unique insights on delineation of post-symptomatic and pre-symptomatic subjects. There also appears to exist a group of entirely asymptomatic subjects who had no sick leave neither before nor after sampling.

Strengths of our study include the fact that it was a large and systematically enrolled cohort that used administrative sick leave data and was therefore not hampered by recall bias to which studies sourcing information from participants can be subjected. Weaknesses of our study include that we were not able to study the relation of biomarkers to infections occurring more than 6-7 weeks before testing, as community transmission of SARS-CoV-2 started in our region only about 6-7 weeks before the study. Also, employees who were not at work were not eligible for inclusion which is likely to have resulted in an underestimation of the spread of the infection at the hospital as employees may have been absent because of COVID-19. The fact that some participants did not have both tests completed is not likely to have affected results, as lack of analysis results was a random phenomenon and the study was still substantially overpowered. Finally, participants were not questioned about present or prior symptoms. The hospital rules were clear that employees with symptoms should not be at work and we had, by design, decided to use only sick leave data to avoid possible recall bias. Subjects may of course have sick leave for many other reasons than Covid-19, but the increases of total sick leave associated with SARS-CoV-2 test positivity was greatly increased compared to the sick leave rates for SARS-CoV-2 negative subjects.

We conclude that the amount of virus as determined by the Ct value of the PCR test and also the serology status are useful testing results for distinction between post-symptomatic, asymptomatic, and pre-symptomatic subjects. This is essential for optimal identification of subjects to be targeted by infection control programs in a phase of the epidemic where many exposed and still positive subjects may have recovered quite some time ago. Prior evidence seems clear that pre-symptomatic subjects are indeed infectious 0-2 days before onset of symptoms and that pre-symptomatic subjects significantly contribute to the spread of the infection [2]. As infectivity declines rapidly after the debut of symptoms, it seems more useful to detect infected subjects before the debut of symptoms, rather than after the symptoms have already been present for some time. When the epidemic has been ongoing for some time, testing strategies need to ascertain whether a test positivity reflects a post-symptomatic infection or whether it may reflect a high risk for a pre-symptomatic infection.

In summary, we propose that systematic SARS-CoV-2 screening of healthy subjects may be useful also in a phase of the epidemic where many positive subjects have had previous disease, as the Ct value of the PCR result may predict if subjects are in a pre-symptomatic phase.

## Supporting information

STROBE

## Data Availability

The data constitutes sensitive data about health of human research subjects. However, pseudonymised, individual-level data that allow full replication of the results in this article will be made freely available from. The study protocol is available at clinicaltrials.gov NCT04411576

## Acknowledgements

We would like to thank Suyesh Amatya, Helena Andersson, Shaghayegh Bayati, Emine Eken, Pedram Farsi, Yasmin Hussein, Roxana Merino Martinez, Sara Mravinacova, Björn Pfeifer, Ulla Rudsander, Ronald Sjöberg, Lovisa Skoglund, Balazs Szakos, Hanna Tegel, Emel Yilmaz and Jamil Yousef for excellent technical assistance.

## Ethical approval

The study was approved by the Swedish Ethical Review Authority (approval number 2020-01620).

