## Supplemental table 1 for "Screening for high amounts of SARS-CoV-2 identifies pre-symptomatic subjects among healthy healthcare workers"

| **Table S1. Bivariate association between covariates and sick leave (not mutually adjusted)** | | | |
| --- | --- | --- | --- |
|  | **1-2 weeks after testing vs No sickleave OR (95% CI)** | **1-3 weeks before testing vs No sickleave OR (95% CI)** | **4-6 weeks before testing vs No sickleave OR (95% CI)** |
| **Age** | | | |
| 20-29 | 1·00 | 1·00 | 1·00 |
| 30-39 | 1·06 (0·79-1·40) | 1·08 (0·88-1·32) | 1·14 (0·96-1·37) |
| 40-49 | 0·61 (0·45-0·83) | 0·89 (0·72-1·09) | 0·93 (0·78-1·12) |
| 50-59 | 0·62 (0·46-0·85) | 0·79 (0·64-0·98) | 0·90 (0·75-1·08) |
| 60+ | 0·46 (0·32-0·66) | 0·62 (0·49-0·78) | 0·77 (0·63-0·94) |
| **Sex** | | | |
| Female | 1·00 | 1·00 | 1·00 |
| Male | 0·61 (0·48-0·77) | 0·54 (0·46-0·64) | 0·60 (0·52-0·69) |
| **SARS-CoV-2 test results** | | | |
| PCR neg/Serology neg | 1·00 | 1·00 | 1·00 |
| PCR neg/Serology pos | 1·04 (0·70-1·56) | 3·59 (2·98-4·33) | 3·34 (2·83-3·95) |
| PCR pos/Serology neg | 7·47 (4·72-11·84) | 1·94 (1·14-3·31) | 1·31 (0·78-2·21) |
| PCR pos/Serology pos | 3·76 (1·60-8·85) | 15·90 (1·80-25·79) | 2·48 (1·32-4·69) |
| **SARS-CoV-2 PCR test results** | | | |
| PCR negative | 1·00 | 1·00 | 1·00 |
| PCR weakly positive | 3·68 (2·08-6·49) | 6·23 (4·35-8·93) | 1·43 (0·89-2·29) |
| PCR strongly positive | 12·58 (6·67-23·73) | 3·69 (1·89-7·19) | 1·80 (0·87-3·72) |
